# Validation of a Smartphone-Image-Based Computer-Vision Model for Lean Mass and Body Fat Estimation Against Dual-Energy X-ray Absorptiometry

**DOI:** 10.64898/2026.06.15.26355736

**Authors:** Tobias Reynolds, Jackson Gerard, Aydan Jordan, Nathan Streby, Brynlee Stoll, Andrew Bowers, Anna Hewitt, Jessica Bargamian, Teryn Sapper, Timothy E. Burdick, Madison Kackley

**Affiliations:** Kino Vision, Inc., West Chicago, Illinois, USA; Department of Human Sciences, The Ohio State University, Columbus, Ohio, USA; Community and Family Medicine, Dartmouth Health, Lebanon, New Hampshire, USA; Biomedical Data Science, Dartmouth College, Hanover, New Hampshire, USA

## Abstract

**Introduction:** Body composition, rather than body weight alone, is an increasingly important health metric, and preservation of lean mass has become a central concern in obesity treatment, aging, and chronic disease management. Dual-energy X-ray absorptiometry (DXA) provides accurate assessment of fat and lean tissue, but its cost and logistical requirements limit repeated measurement. Computer-vision approaches show promise for estimating adiposity from smartphone images, but lean-mass estimation remains less established.

**Methods:** We evaluated a computer-vision body composition model, applied to consumer-grade smartphone photographs, against DXA in a held-out validation sample of 195 adults from an ongoing cross-sectional study. Body fat percentage and total lean mass percentage were co-primary outcomes; for total lean mass percentage, an image-only configuration (no added covariates) was pre-specified as primary. Agreement was quantified using Lin’s concordance correlation coefficient (CCC) as the lead statistic, with Pearson correlation, mean absolute error, root mean square error, mean bias, and Bland-Altman limits of agreement. In secondary analyses, appendicular lean mass and total lean mass percentage were each estimated with and without routine anthropometric and demographic inputs (body weight, height, age, and sex).

**Results:** Total lean mass percentage agreed with DXA from image features alone (CCC 0.916). Body fat percentage, estimated with routine inputs added, agreed at least as closely (CCC 0.930). Adding routine inputs barely changed agreement for total lean mass percentage but markedly improved it for appendicular lean mass, an absolute quantity that scales with body size.

**Conclusions:** A smartphone-image-based model estimated both body fat and lean mass with strong agreement to DXA, with lean mass percentage from image features alone. The approach needs no fixed equipment or ionizing radiation. Whether it can track change over time, including in incretin-based weight loss where lean mass preservation is a concern, was not assessed in this cross-sectional study.

**Author summary:** Fat and muscle content in a person explains more about their health than weight alone, but the most accurate way to measure body composition requires specialized scanning equipment that is vastly inaccessible. We tested whether smartphone photographs could be used in an artificial-intelligence model to estimate body fat and muscle in a way that agrees with such medical scans. In a group of 195 adults, estimates from smartphone photographs agreed closely with the scan for both body fat and muscle. Measuring muscle this way is the more novel result, because earlier smartphone studies focused mostly on body fat, and monitoring muscle loss is becoming increasingly important with newer diabetes and weight-loss medications. We also looked at how much a few simple pieces of routine information, a person’s weight, height, age, and sex, changed the estimates. Adding this information made little difference when we expressed muscle as a share of the body, but helped substantially when we expressed muscle as an absolute amount, because absolute muscle tracks closely with overall body size. These results suggest a smartphone could make body composition easier to measure regularly, though more research is needed in larger and more diverse groups before it can become standard.

## Introduction

Body composition is a more clinically meaningful marker of health for a range of conditions than conventional anthropometric measures such as body mass index (BMI), because BMI cannot distinguish fat from lean tissue and performs inconsistently across sex, age, and ancestry [1]. This distinction matters in disease management as well as wellness. Lean mass in particular has become a focus of clinical attention. In the treatment of type 2 diabetes with incretin-based therapies such as glucagon-like peptide-1 (GLP-1) and glucose-dependent insulinotropic polypeptide (GIP) receptor agonists, substantial weight loss can be accompanied by loss of lean mass, and preservation of skeletal muscle has become a central concern in evaluating these therapies [2]. The ability to measure lean mass accessibly, rather than relying only on weight, BMI, or adiposity, is therefore becoming increasingly relevant to clinical care.

Dual-energy X-ray absorptiometry (DXA) is widely used as a reference method for body composition because it provides accurate, regional estimates of fat mass, lean soft-tissue mass, and bone mineral content [3]. Its use outside research and specialty settings is limited by cost, fixed equipment, a trained-operator requirement, and ionizing radiation. These constraints have motivated interest in methods that approximate DXA-derived body composition without specialized hardware and that are accessible enough to support frequent measurement over time.

Smartphone-based imaging is one such approach. Body fat percentage estimated from smartphone photographs has shown strong agreement with DXA in controlled validation studies [4,5], and smartphone-derived anthropometric features have been used to estimate appendicular lean mass and skeletal muscle [6]. Other smartphone-based approaches to body composition, including bioelectrical impedance, have also been explored [7]. Review articles describe a growing set of mobile applications with varying methods, reference standards, and reported accuracy [8]. Lean-mass estimation in particular remains less well characterized than adiposity despite its growing clinical importance, and estimating lean mass directly from images, rather than inferring it from extracted anthropometric features, has received little attention.

A central question for image-based lean-mass estimation is how much of a model’s accuracy derives from the image itself versus from routine inputs such as body weight, height, age, and sex, which are themselves strong predictors of lean mass. This distinction matters because a model can appear accurate while drawing most of its predictive information from these inputs rather than from the photograph. Prior work combining smartphone images with additional non-image inputs has not isolated the independent contribution of each [9]. The contribution of these inputs may also depend on the estimation target, since absolute lean mass and lean mass expressed as a percentage relate differently to overall body size. We therefore evaluate lean-mass estimation under two input conditions, image-derived features alone and image-derived features combined with routine anthropometric and demographic inputs, to test whether these inputs contribute differently across targets.

In this study, we evaluate a smartphone-image-based body composition model against DXA in a held-out validation sample drawn from an ongoing cross-sectional study. Agreement for body fat percentage and total lean mass percentage are co-primary validation outcomes. As secondary analyses, we evaluate appendicular lean mass and total lean mass percentage under the two input conditions described above to characterize the contribution of routine anthropometric and demographic inputs across estimation targets. We hypothesized that smartphone-derived body fat percentage and total lean mass percentage would each show strong agreement with DXA, and that the added value of these inputs would be greater for absolute lean mass than for lean mass expressed as a percentage. This validation is the first in a planned program of work establishing the platform’s measurement performance across body composition targets and populations.

## Materials and methods

### Study design and oversight

We report a planned validation analysis of body composition agreement from an ongoing cross-sectional study (The Ohio State University IRB-approved protocol STUDY20250275) comparing body composition estimates derived from consumer-grade smartphone photographs against DXA as the reference standard. All participants provided written informed consent before any study procedure. The body composition model under evaluation, Vision Body Composition Plus (VBC+; Kino Vision, Inc.), was used solely for research correlation purposes; participants did not receive model-generated results, and no VBC+ output was used to inform any clinical decision-making.

### Participants

Eligible participants were adults aged 18 to 75 years who were capable of providing informed consent and willing to undergo DXA, anthropometric measurement, and standardized smartphone photography. Exclusion criteria were pregnancy (verified by urine human chorionic gonadotropin test for participants who could become pregnant), the presence of an implanted medical device (e.g., a pacemaker), and major surgery or serious injury within the preceding six months. Participants were recruited through IRB-approved flyers, university listservs, and social media. Eligibility was confirmed via a brief intake questionnaire at enrollment.

Data collection for the parent study is ongoing. The validation set analyzed here comprised the 195 participants enrolled through the data cutoff. No eligible participant was excluded after enrollment; all 195 participants completed both a DXA scan and standardized smartphone photography with usable data and were included in every agreement analysis.

### Reference Standard

Whole-body composition was assessed by DXA using a GE Lunar iDXA (GE HealthCare) with enCORE software version 14.10. Scans were acquired by trained, certified study personnel following the manufacturer’s standardized procedures and were analyzed independently of the study team members affiliated with the model developer (see Competing Interests). DXA-derived body fat percentage, total lean mass percentage, and appendicular lean mass served as reference values. Smartphone image capture and DXA were performed within 15 minutes of one another to ensure that reference measurements reflected body composition at the time of imaging.

### Anthropometric Measurements

Waist and hip circumferences were measured by trained study personnel using a flexible anthropometric tape, obtained in duplicate at the narrowest point of the torso and the widest point of the hips, respectively, with the mean recorded. These circumference measurements were collected per the study protocol but were not used as inputs to any VBC+ configuration evaluated in this analysis; the model inputs comprised body weight, height, age, and sex.

### Smartphone image acquisition

Participants were photographed in a private laboratory setting by trained staff using a standardized four-view protocol, comprising anterior, posterior, and bilateral lateral views. Participants stood in a defined standing position with feet approximately shoulder-width apart and arms held slightly away from the torso to reduce limb occlusion. Male participants were photographed without a shirt, wearing shorts; female participants wore a sports bra and shorts. A single image set was captured per participant. Images were captured using the front-facing cameras of multiple consumer devices (Apple iPhone 14, iPhone 15, and 4th-generation iPad Pro) to introduce device variability representative of real-world capture; front-facing cameras were used to reflect the intended self-capture use case, in which a user can frame themselves without assistance from a second person. Images were acquired across multiple laboratory rooms under varying ambient lighting conditions to reflect realistic capture variability.

### Image processing and de-identification

Captured images were preprocessed to isolate the participant from the image background prior to analysis. Participant faces were digitally blurred using open-source computer-vision software before storage and analysis, in accordance with the approved protocol; no unblurred facial images were used in this analysis. Study data were maintained in a secure, access-controlled institutional REDCap database hosted by The Ohio State University, with participant identity protected through de-identification and access restricted to authorized study personnel.

### Body composition model

The model evaluated in this study is a machine-learning (computer-vision) system, not a computation performed on the phone: the smartphone serves as the image-capture device, and background-removed images are analyzed on cloud infrastructure. Background-removed images were provided to VBC+ (Vision Body Composition+; the research designation of Kino Vision’s proprietary KINO model), a convolutional neural network that generates body composition estimates directly from image data, with participant body weight, height, age, and sex incorporated as additional inputs to refine predictions in the deployed configuration. The model produced estimates of body fat percentage, total lean mass percentage, and appendicular lean mass.

### Model development data

The model was developed on a separate dataset of approximately 1,050 paired smartphone-image and DXA records, collected independently of the validation study reported here. Reference labels for development were obtained by DXA scans performed by trained personnel following standardized whole-body procedures. Validation records were flagged and stored separately at the point of collection and excluded from all model development so that no participant in the 195-person validation set contributed to training; development and validation sets are disjoint at the participant level. Development data spanned a range of body types and demographic backgrounds broadly reflecting the U.S. adult population; detailed demographic composition of the development set is available from the authors upon reasonable request. Specific model architecture and training procedures are proprietary; this study evaluates the agreement of the deployed model’s outputs against DXA on held-out data and does not require disclosure of model internals.

### Statistical analysis

The primary objective was to quantify agreement between smartphone-derived estimates and DXA for two co-primary outcomes: body fat percentage and total lean mass percentage. For total lean mass percentage, the image-only configuration (image-derived features without any added covariates) was designated the primary configuration; the comparison of image-only and covariate-augmented estimation is reported as a secondary analysis.

Each outcome was assigned its primary configuration in advance based on principled grounds rather than by post hoc selection. For the body-size-independent percentages, routine inputs such as weight, height, age, and sex carry little information because the outcome is already independent of body size. Total lean mass percentage was therefore reported image-only, which isolates what the photograph alone contributes. Body fat percentage was reported in the deployed image-plus-inputs configuration, consistent with prior smartphone adiposity validation [4,5] and matching how the model is used; because image-based adiposity estimation is already well validated [4,5], an image-only body fat configuration was not a reporting target. Each co-primary outcome is thus reported in the configuration that fits the question it answers. The absolute lean-mass targets (appendicular and total lean mass in pounds) scale directly with body size, so both were reported under both configurations. Specifying these analyses in advance prevents configuration selection based on observed agreement.

For each co-primary outcome, we computed Lin’s concordance correlation coefficient (CCC) as the primary agreement statistic, together with the Pearson correlation coefficient, mean absolute error (MAE), root mean square error (RMSE), and mean bias. The 95% confidence interval for each CCC was derived via the inverse hyperbolic tangent (Fisher Z) transformation. Agreement was further characterized by Bland-Altman analysis with 95% limits of agreement. CCC was designated the lead statistic because it reflects both precision and systematic deviation, whereas correlation alone does not penalize bias.

As secondary analyses, appendicular lean mass (in absolute units) and total lean mass percentage were each estimated under two conditions: image-derived features alone, and those features combined with body weight, height, age, and sex. This allowed the ability to gauge how much the non-image inputs added to lean-mass estimation.

Pre-specified exploratory analyses examined (i) whether the contribution of these added inputs to lean-mass estimation differs between absolute appendicular lean mass and total lean mass percentage, and (ii) agreement for body fat percentage stratified by sex and adiposity category (lean, average, higher). Stratified analyses were descriptive, and the lowest-adiposity stratum was excluded a priori owing to insufficient sample size. The validation sample size was determined by the primary objective of estimating agreement with adequate precision rather than by hypothesis-test power. A sample of approximately 195 participants was projected to estimate the concordance correlation coefficient for each co-primary outcome with a 95% confidence interval half-width of approximately 0.02 to 0.03 at anticipated CCC values above 0.85, which we judged sufficient to characterize agreement at the group level. Subgroup analyses were not separately powered, and stratified estimates are reported as descriptive. Analyses were performed in Python 3.11 using NumPy, pandas, and SciPy; figures were generated with Matplotlib.

## Results

A total of 195 participants comprised the held-out validation set (Table 1). Participants had a mean age of 34.2 years (range 18 to 75) and were approximately evenly split by sex (100 male, 51.3%; 95 female, 48.7%). Mean BMI was 26.8 kg/m². DXA-derived reference values spanned a wide range of body composition, with body fat percentage from 7.5 to 50.6% (mean 27.8%), total lean mass percentage from 47.6 to 89.3% (mean 69.0%), and appendicular lean mass from 29.9 to 105.3 pounds (mean 57.2). The cohort was predominantly white (73.8%) and university-affiliated, with limited representation of some racial, ethnic, and older-age groups, consistent with the single-site recruitment described in the Methods.

**Table 1.** Cohort characteristics and DXA-derived reference values (n = 195).

| Characteristic | Value |
| --- | --- |
| n | 195 |
| Age (years), mean ± SD (range) | 34.2 ± 13.6 (18-75) |
| **Sex** |  |
| Male | 100 (51.3%) |
| Female | 95 (48.7%) |
| **Race/ethnicity** |  |
| White | 144 (73.8%) |
| East Asian | 15 (7.7%) |
| Black/African American | 14 (7.2%) |
| South Asian | 11 (5.6%) |
| Other/multiracial | 7 (3.6%) |
| Hispanic/Latino | 3 (1.5%) |
| Not reported | 1 (0.5%) |
| BMI (kg/m<sup>2</sup>), mean ± SD | 26.8 ± 4.6 |
| **DXA reference measurements** |  |
| Body fat (%), mean ± SD (range) | 27.8 ± 9.2 (7.5-50.6) |
| Total lean mass (%), mean ± SD (range) | 69.0 ± 8.9 (47.6-89.3) |
| Total lean mass (lbs), mean ± SD (range) | 119.9 ± 29.8 (69.9-212.9) |
| Appendicular lean mass (lbs), mean ± SD (range) | 57.2 ± 16.3 (29.9-105.3) |

### Lean mass

For total lean mass percentage, the pre-specified primary configuration used image-derived features without any added covariates (n = 195). Agreement with DXA was strong, with a CCC of 0.916 (95% CI 0.894 to 0.933) and a Pearson correlation of 0.920. MAE was 2.85 percentage points, and RMSE was 3.55 percentage points. The mean bias was −0.60 percentage points, with 95% limits of agreement from −7.47 to +6.28 percentage points (Fig 1). The predicted-versus-reference relationship is shown in Fig 4. Estimating lean mass as a proportion of body composition from image features alone extends prior smartphone-based work that has focused predominantly on adiposity. Agreement statistics for both co-primary outcomes are reported in Table 2.

**Fig 1.**
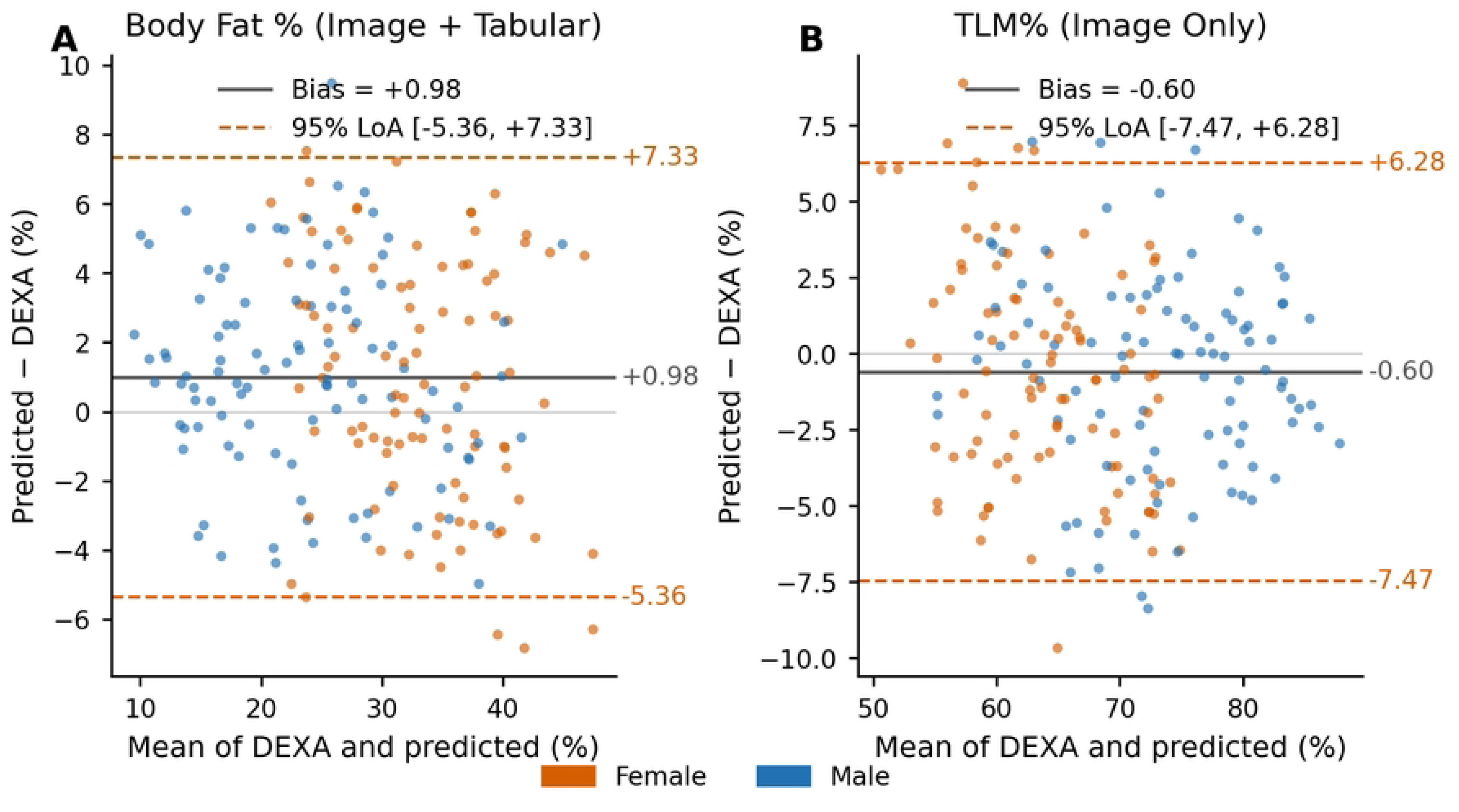
Bland-Altman plots of agreement between smartphone-derived estimates and DXA for the co-primary outcomes. Panel A, body fat percentage (image plus inputs); panel B, total lean mass percentage (image only). Solid line, mean bias; dashed lines, 95% limits of agreement. Points are colored by sex.

**Table 2.** Agreement between smartphone-derived estimates and DXA for the co-primary outcomes (n = 195).

| Outcome (configuration) | n | CCC (95% CI) | Pearson r | MAE | RMSE | Bias |
| --- | --- | --- | --- | --- | --- | --- |
| Total lean mass % (image only) | 195 | 0.916 (0.894-0.933) | 0.920 | 2.85 | 3.55 | -0.60 |
| Body fat % (image + inputs) | 195 | 0.930 (0.911-0.943) | 0.937 | 2.77 | 3.38 | +0.98 |

CCC, Lin's concordance correlation coefficient; CI, confidence interval; MAE, mean absolute error; RMSE, root mean square error. Errors and bias for body fat and total lean mass percentage are in percentage points. Total lean mass percentage is shown in the pre-specified image-only primary configuration; body fat percentage was estimated in the deployed image-plus-inputs configuration.

### Body fat percentage

Smartphone-derived body fat percentage showed strong agreement with DXA across the validation set (n = 195). The CCC was 0.930 (95% CI 0.911 to 0.943) with a Pearson correlation of 0.937. MAE was 2.77 percentage points, and RMSE was 3.38 percentage points. Bland-Altman analysis showed a small positive mean bias of 0.98 percentage points, with 95% limits of agreement from −5.36 to +7.33 percentage points (Fig 1).

### Contribution of routine inputs by estimation target

To see how much the routine inputs add, we estimated total lean mass percentage and appendicular lean mass under two input conditions: image-derived features alone and image-derived features plus body weight, height, age, and sex (Table 3).

**Table 3.** Lean-mass agreement under two input conditions: image-derived features alone versus image-derived features combined with routine anthropometric and demographic inputs (body weight, height, age, and sex).

| Outcome / input condition | n | CCC (95% CI) | Pearson r | MAE | RMSE | Bias | 95% LoA |
| --- | --- | --- | --- | --- | --- | --- | --- |
| Total lean mass % : image only | 195 | 0.916 (0.894-0.933) | 0.920 | 2.85 | 3.55 | -0.60 | -7.47 to +6.28 |
| Total lean mass % : image + inputs | 195 | 0.923 (0.902-0.939) | 0.927 | 2.75 | 3.39 | -0.52 | -7.11 to +6.07 |
| Appendicular lean mass (lbs) : image only | 195 | 0.881 (0.851-0.907) | 0.903 | 5.75 | 7.33 | +1.82 | -12.13 to +15.78 |
| Appendicular lean mass (lbs) : image + inputs | 195 | 0.970 (0.962-0.977) | 0.972 | 3.02 | 3.88 | +0.43 | -7.14 to +8.00 |
| Total lean mass (lbs) : image only | 195 | 0.888 (0.859-0.912) | 0.904 | 10.03 | 13.04 | +1.89 | -23.47 to +27.24 |
| Total lean mass (lbs) : image + inputs | 195 | 0.970 (0.960-0.977) | 0.973 | 5.31 | 7.07 | -1.38 | -15.00 to +12.23 |

CCC, Lin's concordance correlation coefficient; CI, confidence interval; MAE, mean absolute error; RMSE, root mean square error; LoA, Bland-Altman 95% limits of agreement. Percentage outcomes are in percentage points; appendicular and total lean mass in absolute pounds. Body weight, height, age, and sex were the inputs added in the image-plus-inputs condition. Body fat percentage is reported only in the deployed image-plus-inputs configuration; an image-only body fat configuration was not a reporting target because image-based adiposity estimation is already well validated [4,5], as pre-specified in the Statistical analysis.

For total lean mass percentage, adding these inputs produced negligible change in agreement: the CCC was essentially unchanged (0.916 image-only vs. 0.923 with inputs), as were MAE (2.85 vs. 2.75 percentage points) and RMSE (3.55 vs. 3.39 percentage points).

The absolute targets behaved differently. For appendicular lean mass, adding these inputs improved agreement substantially: the CCC rose from 0.881 (95% CI 0.851 to 0.907) to 0.970 (95% CI 0.962 to 0.977), MAE decreased from 5.75 to 3.02 pounds, and RMSE decreased from 7.33 to 3.88 pounds. Bland-Altman analysis showed that the added inputs reduced both bias and dispersion: mean bias moved from +1.82 pounds to +0.43 pounds, and the 95% limits of agreement narrowed from −12.13 to +15.78 pounds to −7.14 to +8.00 pounds. The corresponding Bland-Altman plots are shown in Fig 2, and the predicted-versus-reference relationship under both configurations is shown in Fig 3.

**Fig 2.**
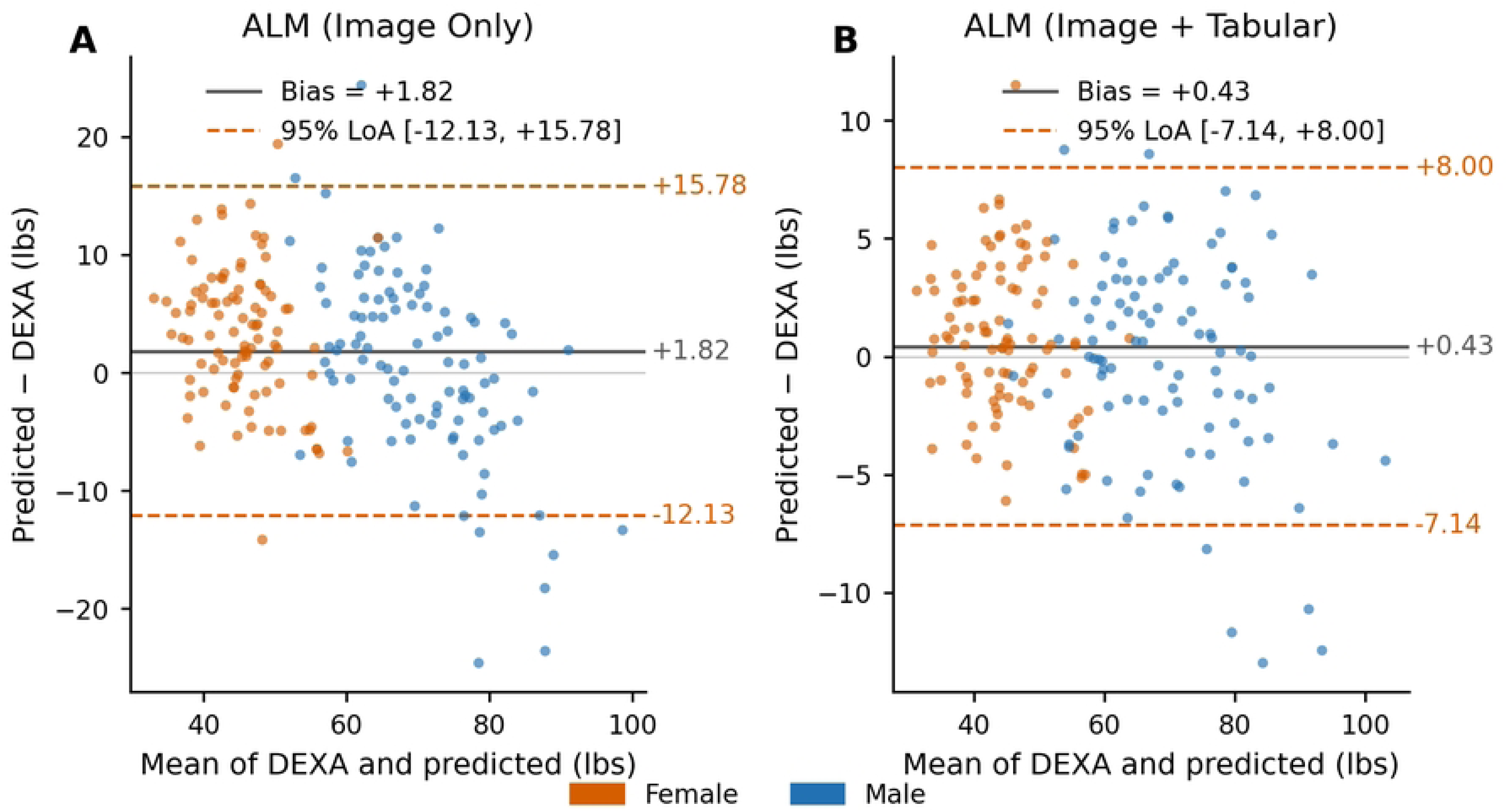
Bland-Altman plots of agreement between smartphone-derived appendicular lean mass and DXA. Panel A, image only; panel B, image plus inputs. Solid line, mean bias; dashed lines, 95% limits of agreement. The added inputs reduce both bias and the width of the limits of agreement. Points are colored by sex.

**Fig 3.**
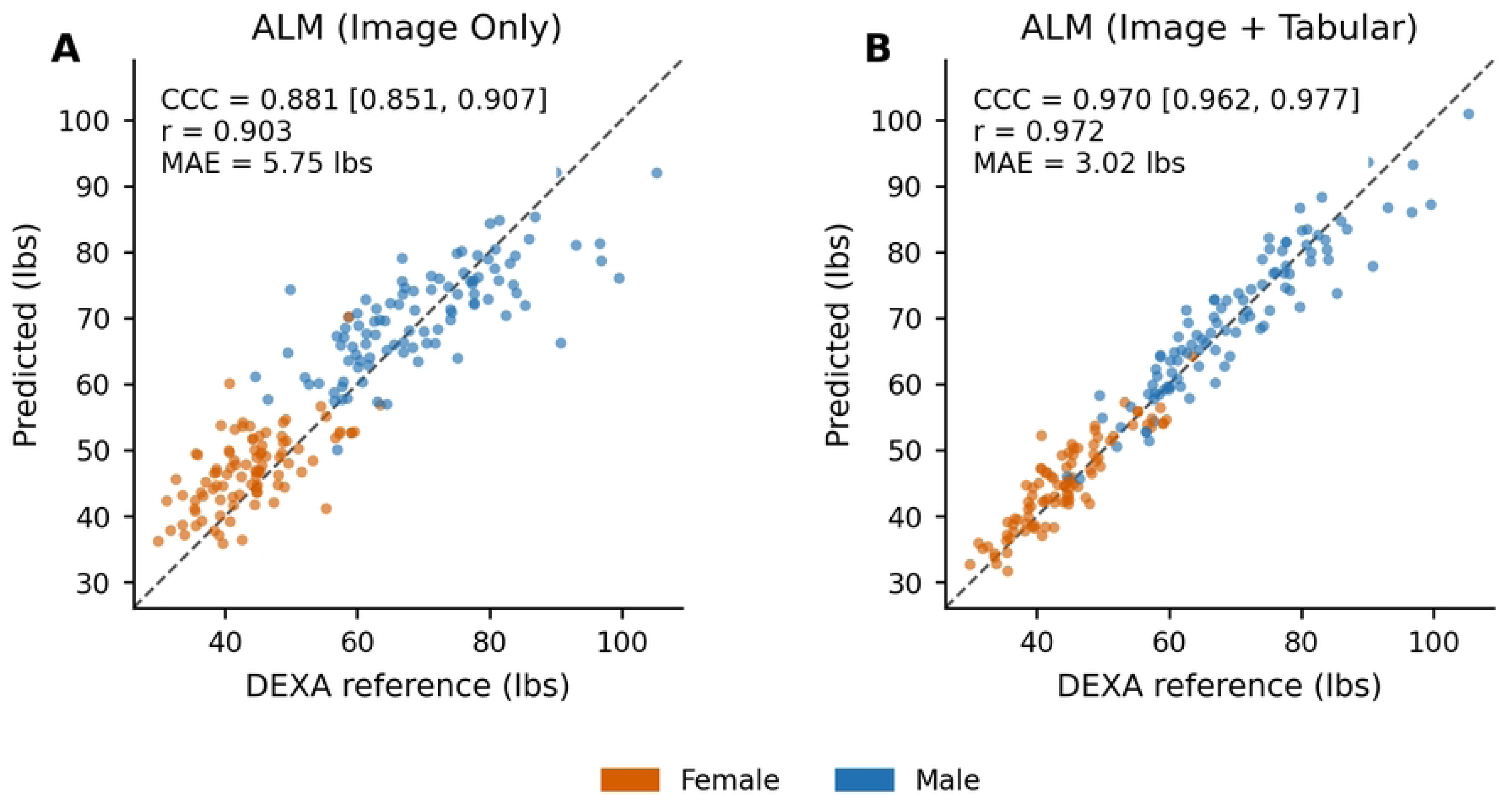
Appendicular lean mass predicted from smartphone images versus DXA reference. Panel A, image only; panel B, image plus inputs. The dashed line is the line of identity. The addition of routine inputs markedly tightens agreement around the line of identity. Points are colored by sex.

**Fig 4.**
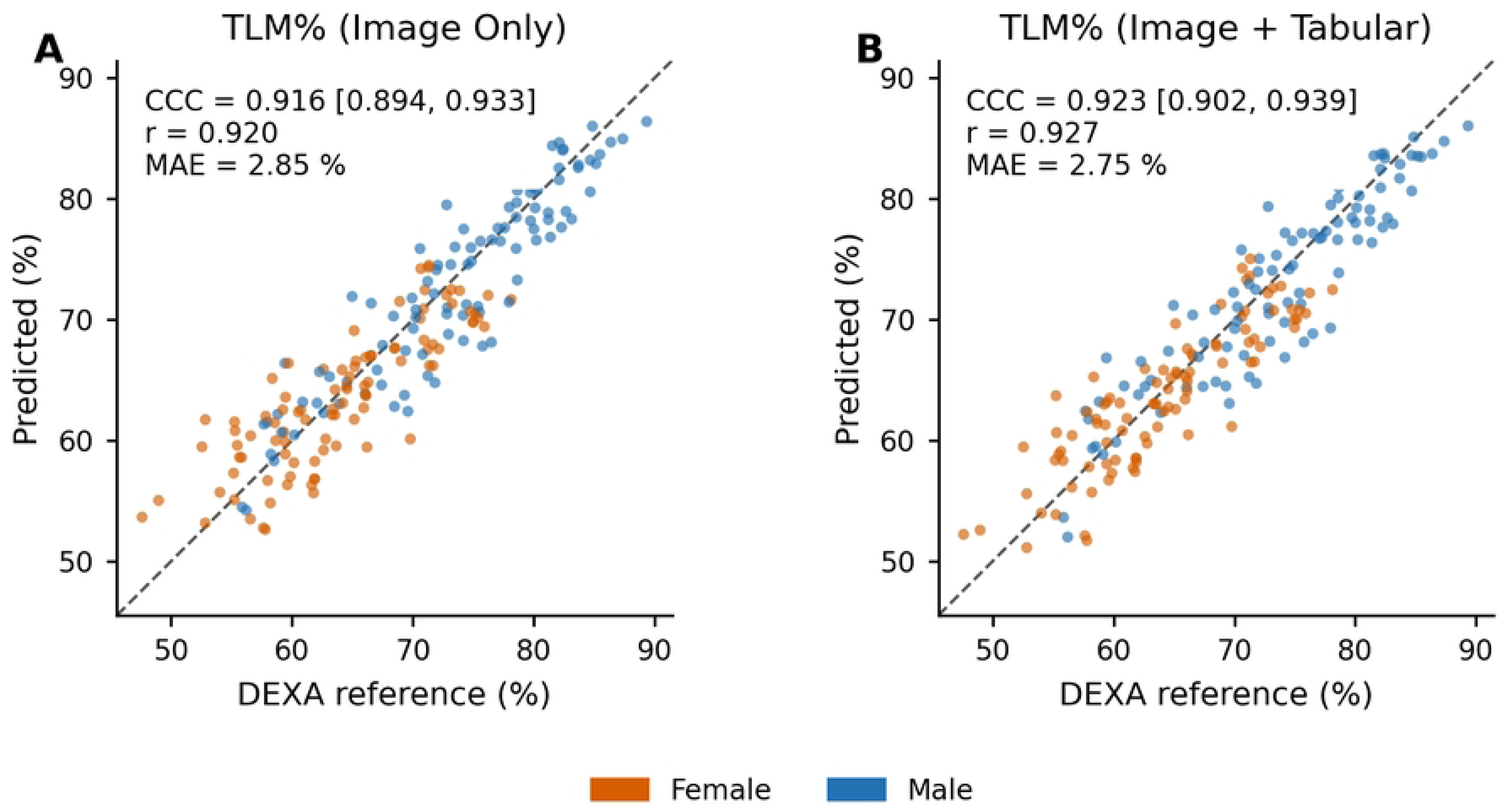
Total lean mass percentage predicted from smartphone images versus DXA reference. Panel A, image only; panel B, image plus inputs. The dashed line is the line of identity. Points are colored by sex.

The same contrast was evident for total lean mass measured in absolute pounds, which we report for completeness alongside the percentage outcome. Image-only estimation yielded a CCC of 0.888 (95% CI 0.859 to 0.912) but wide individual-level error (MAE 10.03 pounds; 95% limits of agreement −23.47 to +27.24 pounds), whereas adding these inputs raised the CCC to 0.970 (95% CI 0.960 to 0.977) and roughly halved the error (MAE 5.31 pounds; 95% limits of agreement −15.00 to +12.23 pounds). Across both absolute lean-mass targets, the added inputs contributed substantially to agreement, while providing little benefit for the body-size-independent percentage outcome.

### Body fat percentage by sex (exploratory)

Agreement for body fat percentage was examined descriptively by sex. Agreement was strong in male participants (CCC 0.933; MAE 2.47 percentage points; n = 100) and somewhat weaker in female participants (CCC 0.852; MAE 3.10 percentage points; n = 95). Within the female participants, agreement was weakest in the lean stratum (BF% < 25%; n = 16), where the model overestimated body fat (MAE 4.47 percentage points; mean bias +4.40), and stronger in the average (n = 45; MAE 2.90) and higher-adiposity (n = 34; MAE 3.49) strata. Among male participants, agreement was strong across all strata: lean (n = 24; MAE 2.11; mean bias +1.91), average (n = 47; MAE 3.15; mean bias +1.22), and higher-adiposity (n = 29; MAE 2.15; mean bias −0.15). Strata were defined for descriptive purposes using approximate sex-specific body fat percentage ranges broadly consistent with widely used fitness categorizations (e.g., American Council on Exercise). The boundary defining the lean female stratum was set higher than the conventional value solely to preserve sufficient cell size for descriptive reporting; this choice was made a priori, before examining agreement within strata, and the strata were not used for any inferential comparison. The lowest-adiposity stratum was excluded a priori owing to insufficient n. These analyses were descriptive and unpowered for subgroup inference; they serve to identify where additional enrollment will most improve performance rather than to estimate subgroup accuracy precisely, and they indicate that the overall sex difference in agreement is concentrated in lean, low-adiposity women.

## Discussion

In a held-out validation sample of 195 adults, a computer-vision model evaluated on smartphone-acquired images showed strong agreement with DXA for both co-primary outcomes. The more novel finding is that the model estimated lean mass, not just adiposity. Total lean mass percentage, derived from image features alone, agreed strongly with DXA, with a CCC above 0.91. Body fat percentage agreed closely as well, with a CCC of 0.93 and an MAE of under three percentage points. Prior smartphone-based work has focused predominantly on adiposity; the present results extend that work to lean mass and show that a single tool can estimate both fat and lean tissue, two compartments that are each clinically informative, from the same set of photographs.

The inputs behaved differently depending on the target. For lean mass as a percentage, adding weight, height, age, and sex changed almost nothing. For lean mass in pounds, the same four inputs improved agreement substantially, whether appendicular or total. This dissociation is interpretable: a body-size-independent ratio carries little information that body-size-correlated inputs can supply, so the image does the work, whereas an absolute mass scales with body size, for which weight, height, and sex are direct proxies. Reporting both configurations guards against a recurring ambiguity in this field, in which a model can appear accurate while drawing its predictive information from routine inputs rather than the photograph. We did not report an image-only body fat configuration. Image-based adiposity estimation has already been validated against DXA in prior work [4,5], so an image-only adiposity arm would have repeated an established result rather than adding new information. The image-only contrast is most informative for lean mass, which is the less characterized target and the focus of this analysis.

These findings sit alongside a small but growing body of smartphone-based validation work. Camera-based adiposity assessment has shown strong agreement with DXA in controlled studies [4,5], and smartphone-derived features have been used to estimate skeletal muscle and appendicular lean mass [6]. Three-dimensional avatar reconstruction from smartphone images has also been applied to body composition assessment, illustrating the range of methods under investigation [10]. A recent scoping review describes a heterogeneous landscape of mobile applications with varying methods, reference standards, and reported accuracy, and notes that lean-mass estimation remains less well characterized than adiposity [8]. The present results contribute to that gap by validating both adiposity and lean-mass percentage against DXA in the same cohort and isolating the role of routine non-image inputs across estimation targets.

The clinical relevance of accessible lean-mass measurement is increasing. In the treatment of type 2 diabetes with incretin-based therapies, substantial weight loss can be accompanied by loss of lean mass, and preservation of skeletal muscle has become a central consideration in evaluating these therapies [2]. Lean-mass loss is likewise a concern in other conditions and treatments, including cancer and cancer-directed therapy [11]. A measurement approach that requires no fixed equipment or ionizing radiation, and that can therefore be repeated frequently, is well suited in principle to tracking change in body composition over time. The present study evaluates measurement agreement at a single time point and does not assess longitudinal tracking, clinical outcomes, or use in any care pathway as those questions are outside its scope. What these results establish is that smartphone-derived estimates of body fat and lean mass agree strongly with a reference method at the group level, which is a necessary prerequisite for future clinical application but not a demonstration of clinical utility.

Several features of the analysis support the credibility of these estimates. The validation set was held out entirely from model development, so the reported agreement reflects performance on data the model did not see. DXA was used as the reference standard, with imaging and scanning performed within a short interval to minimize the likelihood of true body composition changes between measurements. Agreement was characterized using concordance correlation as the lead statistic, which penalizes both imprecision and systematic deviation, and Bland-Altman limits of agreement, which describe error at the level of the individual rather than the group. Reporting both metrics is important: as the absolute lean-mass results illustrate, a high concordance correlation can coexist with individual-level limits of agreement wide enough to matter for applications that depend on a single measurement. We therefore report both throughout so that group-level and individual-level performance can be judged separately.

### Limitations

This study has several limitations, each pointing to a defined next step in an ongoing program of work.

First, the validation sample analyzed here was drawn from a single site. The held-out validation cohort reflects one recruitment setting and a predominantly university-affiliated population. Representation of some racial, ethnic, and age groups was limited in this cohort, which constrains generalizability. Enrollment is ongoing and is expanding representation across these groups, and subsequent analyses will report agreement in a more demographically balanced validation sample.

Second, this analysis was cross-sectional. Each participant contributed a single image set, so the repeatability of estimates across multiple captures was not assessed. Because tracking change over time is an anticipated application, test-retest reliability is a planned focus of subsequent work and should be characterized directly rather than inferred from cross-sectional agreement. Relatedly, image capture was limited to three Apple device models; generalizability across the broader range of smartphone cameras, including Android devices and older sensors, was not assessed and is a target for subsequent work. Separately, reference labels for model development were acquired at a different facility from the validation scans. Although all scans used DXA with standardized whole-body procedures, differences in scanner instrumentation between development and validation could introduce systematic differences in reference values; the strong agreement observed on held-out validation data suggests any such effect was limited.

Third, agreement for body fat percentage was strong overall but weaker in female participants than in male participants (CCC 0.852 vs. 0.933). Part of this gap reflects relative scale: at the lower end of the adiposity range, where female participants are more represented, a given absolute error in percentage points corresponds to a larger fractional error because it is referenced against a smaller body fat percentage. These comparisons were descriptive and unpowered for subgroup inference; they serve to identify where additional enrollment will most improve performance rather than to estimate subgroup accuracy precisely.

Fourth, lean-mass estimates that combine image-derived features with routine anthropometric and demographic inputs reflect a deployable configuration rather than the independent contribution of the image alone. We report the image-only configuration alongside it to keep this distinction explicit so that the input-supported estimates are not misread as image-derived performance. The input-supported condition combined four inputs, and the present analysis does not decompose their individual contributions. The relative role of body weight, height, age, and sex is a target for future ablation work.

Finally, this analysis evaluated a single deployed version of the model against DXA. DXA is a reference method rather than a true criterion standard and carries its own measurement error, which sets a practical ceiling on the agreement any comparison method can demonstrate. The results characterize the model version evaluated and are not a claim about subsequent versions, which continue to be developed and validated.

## Conclusion

In a held-out validation sample, a smartphone-image-based model estimated body fat percentage and total lean mass percentage with strong agreement to DXA, and did so for total lean mass percentage from image-derived features alone. The added inputs (weight, height, age, sex) mattered only for the absolute targets: they did little for a body-size-independent ratio but substantially improved agreement for appendicular lean mass. These findings support smartphone-image-based estimation as a measurement approach for both body fat and lean mass and motivate the next steps in this program of work: validation in larger and more demographically diverse samples and direct characterization of test-retest reliability to support tracking change over time.

## Acknowledgments

The authors thank Caulen Spangler, Lucas Winnestaffer, Jake Lagando, Sreyan Banerji, and Gabby Huxtable for their contributions to the study design, operations, and coordination. The authors also thank Siddhartha Chandra for informal scientific discussion on machine-learning approaches. The concept of estimating body composition from a photograph, and early development of the study protocol by T.R. and J.G., predate the formation of Kino Vision, Inc.; the company was subsequently formed to develop the technology. Competing interests and funding are detailed in the dedicated statements below.

## Data availability

The de-identified participant-level data underlying the agreement analyses (paired model estimates and DXA reference values) contain image-derived biometric information and are subject to the data use terms of The Ohio State University IRB-approved protocol STUDY20250275. They cannot be made publicly available without restriction. De-identified data sufficient to reproduce the reported analyses are available to qualified researchers under a data use agreement administered by The Ohio State University; requests may be directed to the Office of Responsible Research Practices or the corresponding author, who will facilitate the institutional request process. The smartphone images themselves cannot be shared, as they constitute identifiable biometric data even after facial blurring.

## Author contributions

Author contributions are reported using the CRediT taxonomy.

Tobias Reynolds: Conceptualization, Methodology, Writing - original draft, Project administration.

Jackson Gerard: Software, Formal analysis, Data curation, Methodology, Writing - review & editing.

Aydan Jordan: Investigation, Project administration, Writing - review & editing.

Nathan Streby: Investigation, Project administration, Writing - review & editing.

Brynlee Stoll: Investigation, Project administration, Writing - review & editing.

Andrew Bowers: Investigation, Project administration, Writing - review & editing.

Anna Hewitt: Investigation, Project administration, Writing - review & editing.

Jessica Bargamian: Methodology, Writing - review & editing.

Teryn Sapper: Methodology, Supervision, Resources, Writing - review & editing.

Timothy E. Burdick: Conceptualization, Writing - review & editing.

Madison Kackley: Methodology, Investigation, Supervision, Resources, Writing - review & editing.

